# Artificial Intelligence-Enhanced Electrocardiography for Detection and Prediction of Hypertrophic Cardiomyopathy across Monogenic and Polygenic Susceptibility

**DOI:** 10.64898/2026.09.12.26362902

**Authors:** Philip M. Croon, Ryan B. Choi, Evangelos K. Oikonomou, Sumukh V. Shankar, Lovedeep S. Dhingra, Nico Bruining, Peter-Paul Zwetsloot, Michelle Michels, Rudolf A. de Boer, Veronika Puchnerová, Jiří Bonaventura, Robert M.A. van der Boon, Sounok Sen, Rohan Khera

## Abstract

**Background:** Cascade screening increasingly identifies carriers of pathogenic or likely pathogenic sarcomere variants at risk for hypertrophic cardiomyopathy (HCM) in whom penetrance is incomplete, and surveillance relies on resource-intensive serial imaging. We evaluated whether a validated artificial intelligence-enhanced electrocardiography (AI-ECG) model identifies the HCM phenotype at first clinical assessment, predicts development of HCM during follow-up, and complements polygenic risk.

**Methods:** We assembled 1,095 genotype-positive (G+) individuals with pathogenic or likely pathogenic sarcomere variants from Yale-New Haven Hospital (n=119), Erasmus MC (n=858), and Motol University Hospital (n=118). At baseline (first clinical assessment), individuals were classified as phenotype-positive (P+) or phenotype-negative (P−). A previously validated AI-ECG model applied to 12-lead ECG images generated an HCM score. The primary outcome was detection of phenotypic positivity at baseline; secondary analyses included manifest HCM (at baseline or during follow-up) and identifying risk of developing future HCM among G+/P−individuals. In 57,007 UK Biobank participants, we assessed whether AI-ECG adds to an established polygenic risk score (PRS).

**Results:** Among 1,095 G+ individuals (median age 46 years [IQR 34–56]; 52.1% female), 808 (73.8%) were P+ at baseline, 56 (5.1%) developed HCM during follow-up, and 231 (21.1%) remained P−. AI-ECG achieved an AUROC of 0.91 (95% CI 0.89–0.93) for P+ at baseline and 0.92 (95% CI 0.90–0.94) for manifest HCM. At a threshold of 0.15, sensitivity was 0.78, specificity 0.89, PPV 0.95, and NPV 0.59. Among G+/P− individuals, higher AI-ECG scores predicted development of HCM (HR 1.55 per 1-SD; 95% CI 1.28–1.88; p<0.001; adjusted HR 1.38; 95% CI 1.11–1.71; p=0.004). In the UK Biobank, individuals with both high AI-ECG and high PRS had 60-fold higher odds of HCM (adjusted OR 60.2; 95% CI 26.5–137.2), versus 15.0 for high AI-ECG alone and 4.1 for high PRS alone.

**Conclusions:** AI-ECG detects the HCM phenotype at baseline in sarcomere variant carriers, predicts development of HCM in G+/P− individuals, and complements PRS in the general population, supporting AI-ECG as a scalable tool to detect HCM and guide surveillance in individuals with monogenic or polygenic susceptibility.

*Clinical Perspective What Is New?:*

- In 1,095 carriers of pathogenic or likely pathogenic sarcomere variants across three international centers, a validated AI-ECG model applied to 12-lead ECG images detected the HCM phenotype at first clinical assessment (AUROC 0.91).
- Among G+/P− individuals, a higher AI-ECG score predicted development of HCM during follow-up (HR 1.55 per 1-SD increase), with consistent performance across sites, sexes, age groups, and sarcomere genes.
- In 57,007 UK Biobank participants, AI-ECG and polygenic risk provided complementary information; individuals with both a high AI-ECG score and a high polygenic risk score had 60-fold higher odds of HCM.

*What Are the Clinical Implications?:*

- AI-ECG applied to standard ECG images is a scalable, low-cost tool to stratify the risk of HCM in carriers of a sarcomere variant.
- The AI-ECG score may help individualize the intensity of imaging surveillance in G+/P−individuals, focusing resources on those at highest risk of developing HCM.
- Combining AI-ECG with polygenic risk may refine identification of high-risk individuals in the general population and might further improve risk stratification.

## Introduction

Hypertrophic cardiomyopathy (HCM) is the most common inherited cardiomyopathy and is associated with substantial morbidity and mortality, including heart failure, arrhythmias, stroke, and sudden cardiac death.^1^ Although HCM is traditionally considered a Mendelian disease, phenotypic expression reflects an interplay of monogenic variants, polygenic risk, and environmental exposures, resulting in incomplete penetrance and heterogeneity in disease onset and severity.^2–8^ Advances in molecular testing have expanded the identification of pathogenic sarcomere variant carriers, including many who have yet to develop overt HCM.^9^ In addition, in a substantial proportion of clinically diagnosed individuals, no causal sarcomere variant is identified, underscoring the contribution of polygenic and non-genetic factors to disease presentation.^10^ Clinical guidelines, therefore, recommend serial imaging surveillance for at-risk individuals to identify manifestations of HCM over time.^5,11^ Yet this strategy is resource-intensive, variably implemented, and not informed by tools to identify those at greatest near-term risk.

Artificial intelligence (AI) has expanded the diagnostic capabilities of the electrocardiogram (ECG), enabling the detection of structural and functional cardiac abnormalities from standard recordings.^12–14^ Several AI-ECG models have demonstrated strong performance for HCM detection, with the first receiving regulatory approval and supporting clinical deployment.^15^ Preliminary evidence suggests that AI-ECG scores track disease activity over time in response to targeted HCM therapies.^16,17^ However, as a general screening tool, the clinical utility of AI-ECG is limited by the low prevalence of HCM in the population and, consequently, a low positive predictive value.^18^ In contrast, carriers of pathogenic sarcomere variants are at substantially elevated risk of HCM. However, in G+/P− individuals, current approaches provide limited individualized risk stratification, leaving clinicians reliant on repeated, resource-intensive serial imaging to detect phenotypic conversion. Hence, there is a clinical need for tools that stratify the risk of developing HCM beyond serial imaging alone.

In this study, we evaluated whether a validated AI-ECG model developed for the cross-sectional detection of HCM identifies the HCM phenotype at baseline and predicts development of HCM in G+ individuals across three international centers. In addition, among UK Biobank participants with whole-genome sequencing and ECGs, we evaluated whether combining PRS with AI-ECG further improves risk stratification.

## Methods

The study was approved by the institutional review boards at all participating centers, and informed consent was waived due to its retrospective design. The data underlying this study cannot be shared publicly due to patient privacy concerns and institutional regulations.

### Study population

We included carriers of pathogenic or likely pathogenic sarcomere variants from three specialized centers. Eligible genes included *MYBPC3*, *MYH7*, *TNNT2*, *TNNI3*, *TPM1*, *MYL2*, *MYL3*, *ACTC1*, and *ACTN2*.

The Yale cohort comprised all individuals with sarcomere gene variants seeking care at the cardiomyopathy referral center at Yale-New Haven Hospital between February 2013 and June 2023. Phenotype status was determined from clinical imaging reports using a locally deployed large language model (LLM); full methodology and validation are described in the Supplementary Materials. All ECGs were collected in their native DICOM format and transformed into ECG images locally for ECG inference. Comorbidities were abstracted using International Classification of Diseases (ICD) codes documented prior to the index ECG date.

The Erasmus MC cohort included individuals enrolled in a prospective cardiomyopathy registry with systematic genetic testing and longitudinal follow-up at Erasmus Medical Center between September 1994 and December 2024. Phenotypes were determined by the treating clinician in accordance with international guidelines. Genetic variants were classified as pathogenic or likely pathogenic by a cardiac geneticist at the time of molecular testing. Like Yale, the ECGs were collected in native DICOM format and rendered as standard ECG images before the model deployment. The registry has been described in more detail in prior reports.^19^

The cohort from Motol University Hospital consisted of individuals enrolled in a registry for inherited cardiac conditions at a tertiary referral center in Prague between September 1999 and December 2025. All participants had at least one 12-lead ECG available for AI-based analysis. Here, the phenotypes were determined by the treating clinician in accordance with international guidelines. Similarly, the consulting geneticist interpreted the genetic test results and classified them as pathogenic or likely pathogenic. ECGs were scanned copies of clinical ECGs and saved as PDFs. The cohort has been described in prior reports.^20,21^

### AI-ECG model

We calculated the AI-ECG HCM score using a previously validated model consisting of an EfficientNet-B3-based convolutional neural network (CNN), trained to detect HCM in 12-lead ECG images. The development, training, and multinational validation of the model have been described previously.^16^ In addition to broad validation across cohorts, the model has previously been shown to be a dynamic marker of treatment response in patients with HCM.^17^ The model takes a 300×300-pixel representation of a 12-lead ECG as input and outputs a continuous score between 0 and 1. The same model architecture and weights were used across all three sites without retraining.

To enable local inference without requiring technical expertise or data sharing, we developed a software module for macOS and Windows that includes the full inference pipeline for quality control and facilitates easy local deployment from either DICOMs containing the raw signal or ECG images in PDF or JPEG format. This pipeline allowed each participating site to generate AI-ECG predictions without technical expertise or sharing patient data, using ECGs in their native format, preserving patient privacy, and enabling multi-center collaboration.

### Outcome definitions

The primary outcome was the HCM phenotype at baseline, defined as each carrier’s first clinical assessment; carriers were classified as phenotype-positive (P+) or phenotype-negative (P−) based on cardiac imaging at baseline. At Yale, each participant’s earliest cardiac imaging study (echocardiography or CMR) was identified, and the closest ECG within a 90-day window was selected as the index ECG, preferring ECGs obtained before imaging. HCM phenotype status was extracted from the corresponding imaging report using an expert-adjudicated, LLM-assisted annotation pipeline; technical methods and validation are described in the Supplementary Methods. At Erasmus MC and Motol University Hospital, the index ECG was the first available ECG, and phenotype status was defined by the treating clinician based on contemporaneous imaging and international diagnostic guidelines. These registries have been described previously.^4,19^

Carriers without imaging evidence of HCM within the first year after baseline were classified as P− and followed longitudinally for development of HCM. Incident HCM was defined as the first HCM-positive imaging occurring after a 365-day blanking window. The secondary outcome was manifest HCM, defined as HCM present at baseline or during follow-up (prevalent or incident). Carriers without HCM during the entire follow-up period remained P−.

### Comparison and additive value of AI-ECG with polygenic risk

To assess whether the AI-ECG HCM score provides information complementary to genetic risk, we conducted a secondary analysis in the UK Biobank (UKB). We identified all available participants with available ECG and whole-genome sequencing data, restricting this analysis to a genetically homogeneous ancestry group because the HCM polygenic score was developed and evaluated primarily in European-ancestry populations. We calculated their PRS based on the genome-wide PGS for HCM (PGS_MTAG) reported by Zheng et al.^19^ We then applied the AI-ECG model to their 12-lead ECGs to derive the AI-ECG HCM score. HCM was ascertained from inpatient ICD-10 codes (I42.1, I42.2), self-reported cardiomyopathy, and death registry data. In addition, among individuals with available cardiac magnetic resonance (CMR) imaging, HCM was also identified according to the guidelines by a maximum left ventricular wall thickness ≥15 mm. Participants were classified into four groups based on pre-specified thresholds: AI-ECG high (score ≥0.15) or low (<0.15), crossed with PRS high (≥80th percentile) or low (<80th percentile). The 80th percentile was selected to define high polygenic risk, consistent with the top-quintile stratification used in developing the PRS.^19^

### Statistical analysis

Discrimination of the AI-ECG model was assessed using the area under the receiver operating characteristic curve (AUROC), with 95% confidence intervals estimated using the DeLong method, for both P+ at baseline and manifest HCM. AUROCs were calculated for each site individually and for the pooled cohort. Classification metrics, including sensitivity, specificity, positive predictive value (PPV), and negative predictive value (NPV), were computed at a prespecified score threshold of 0.15 for the pooled cohort. Additional thresholds (0.05, 0.10, 0.20, 0.25, 0.30, and 0.50) were evaluated in supplementary analyses. AI-ECG scores were standardized within each site (z-score transformation) for all regression analyses.

To evaluate the prognostic value of the AI-ECG score for HCM development during follow-up, univariate Cox proportional hazards regression models were fitted at each site and in the pooled cohort, with hazard ratios (HRs) reported per 1-standard deviation (SD) increase in the standardized AI-ECG score. Multivariable models were additionally adjusted for age and sex, and the pooled model was further adjusted for site. Model discrimination was assessed using Harrell’s C-index. Patients with zero follow-up time were excluded from Cox regression analyses. For time-to-event analyses, follow-up ended at the earliest of HCM development, initiation of HCM-specific therapy, death, last known follow-up, or a 10-year administrative cap. Site-specific heterogeneity was assessed using Cochran’s Q and I².

Kaplan-Meier survival analysis was performed to estimate phenotype-free survival, stratifying patients into high-risk (AI-ECG score ≥0.05) and low-risk (AI-ECG score <0.05) groups. A lower threshold of 0.05 was used for Kaplan-Meier stratification to maximize sensitivity for early phenotypic signals in G+/P− individuals at baseline under surveillance, whereas the pre-specified threshold of 0.15 was used for phenotype classification in the UK Biobank analysis. Follow-up was capped at 10 years, and survival curves were compared using the log-rank test.

As a secondary analysis, Kaplan-Meier curves were also constructed using patient age as the harmonized time scale, with delayed entry at each patient’s index ECG age. This analysis evaluated age-dependent HCM development during follow-up among G+/P− individuals at baseline under surveillance. A Cox proportional hazards model with age as the time scale was used to compare AI-ECG risk groups. The x-axis was capped at age 60 years due to diminishing numbers at risk beyond that age.

Subgroup analyses were conducted by sex (male, female) and age group (<45, ≥45 years) in the pooled cohort. For each subgroup, AUROC and unadjusted and age-and sex-adjusted Cox hazard ratios per 1-SD increase were computed. Gene-stratified analyses were performed for MYBPC3, MYH7, and a composite of other genes. As a sensitivity analysis, age-stratified subgroup analyses were repeated using age 40 as an alternative cutoff.

For the PRS comparison in the UK Biobank, odds ratios for HCM across the four combined AI-ECG and PRS risk strata were estimated using logistic regression adjusted for age and sex, with the “both low” group (AI-ECG <0.15 and PRS <80th percentile) as the reference category. To test whether the AI-ECG score and PRS captured independent risk information, a logistic regression model was fitted that included the continuous AI-ECG score, continuous PRS (z-score), their interaction term, age, and sex.

## Results

### Study population for monogenic variant analyses

A total of 1,095 individuals with pathogenic or likely pathogenic sarcomere gene variants were included from three centers: 119 from Yale, 858 from Erasmus MC, and 118 from Prague. The median age was 46 years (IQR 34–56), and 570 (52.1%) were female **(Table 1)**. The most common gene variants were MYBPC3 (n=751, 68.6%) and MYH7 (n=163, 14.9%). Hypertension was present in 225 (20.5%) and diabetes in 59 (5.4%).

**Table 1.**
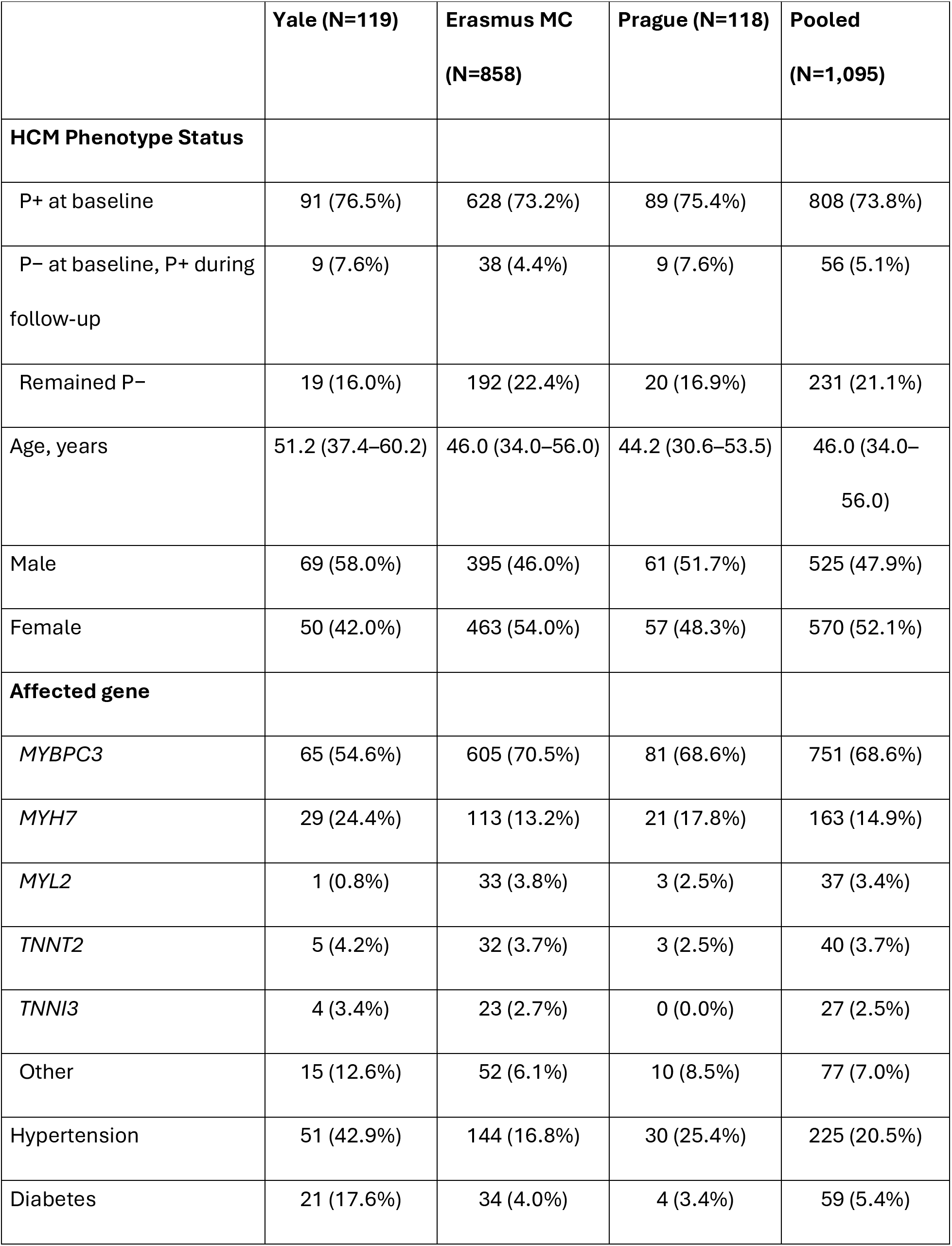
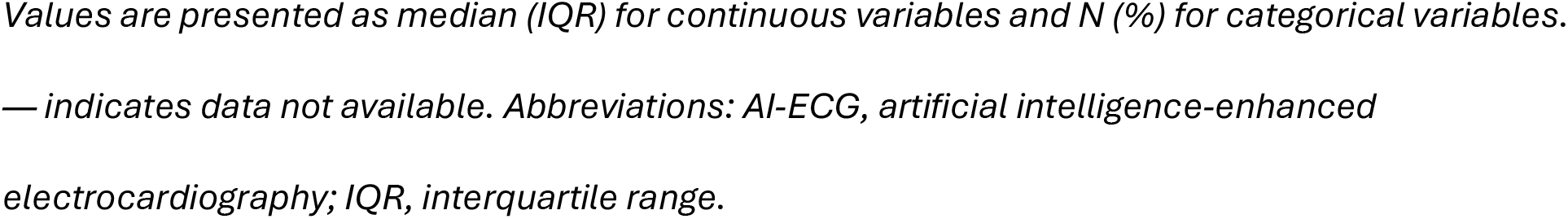
Baseline characteristics of the study population.

|  | Yale (N=119) | Erasmus MC<br>(N=858) | Prague (N=118) | Pooled<br>(N=1,095) |
| --- | --- | --- | --- | --- |
| **HCM Phenotype Status** |  |  |  |  |
| P+ at baseline | 91 (76.5%) | 628 (73.2%) | 89 (75.4%) | 808 (73.8%) |
| P- at baseline, P+ during<br>follow-up | 9 (7.6%) | 38 (4.4%) | 9 (7.6%) | 56 (5.1%) |
| Remained P- | 19 (16.0%) | 192 (22.4%) | 20 (16.9%) | 231 (21.1%) |
| Age, years | 51.2 (37.4–60.2) | 46.0 (34.0–56.0) | 44.2 (30.6–53.5) | 46.0 (34.0–<br>56.0) |
| Male | 69 (58.0%) | 395 (46.0%) | 61 (51.7%) | 525 (47.9%) |
| Female | 50 (42.0%) | 463 (54.0%) | 57 (48.3%) | 570 (52.1%) |
| **Affected gene** |  |  |  |  |
| *MYBPC3* | 65 (54.6%) | 605 (70.5%) | 81 (68.6%) | 751 (68.6%) |
| *MYH7* | 29 (24.4%) | 113 (13.2%) | 21 (17.8%) | 163 (14.9%) |
| *MYL2* | 1 (0.8%) | 33 (3.8%) | 3 (2.5%) | 37 (3.4%) |
| *TNNT2* | 5 (4.2%) | 32 (3.7%) | 3 (2.5%) | 40 (3.7%) |
| *TNNI3* | 4 (3.4%) | 23 (2.7%) | 0 (0.0%) | 27 (2.5%) |
| Other | 15 (12.6%) | 52 (6.1%) | 10 (8.5%) | 77 (7.0%) |
| Hypertension | 51 (42.9%) | 144 (16.8%) | 30 (25.4%) | 225 (20.5%) |
| Diabetes | 21 (17.6%) | 34 (4.0%) | 4 (3.4%) | 59 (5.4%) |

Values are presented as median (IQR) for continuous variables and N (%) for categorical variables. — indicates data not available. Abbreviations: AI-ECG, artificial intelligence-enhanced electrocardiography; IQR, interquartile range.

Of the total cohort, 808 (73.8%) were P+ at baseline, 56 (5.1%) developed HCM during follow-up, and 231 (21.1%) remained P− throughout follow-up. The proportion of G+/P+ individuals at baseline was similar across sites: 76.5% at Yale, 73.2% at Erasmus MC, and 75.4% at Prague. Characteristics at cohort initiation stratified by phenotype group are presented in **Supplementary Table S1**.

### AI-ECG discrimination for P+ at baseline

The AI-ECG model showed strong discrimination for P+ at baseline, with an AUROC of 0.91 (95% CI: 0.89–0.93) in the pooled cohort **(Figure 1, Supplementary Figure S1)**. Site-specific AUROCs were 0.88 (95% CI: 0.81–0.95) for Yale, 0.92 (95% CI: 0.90–0.94) for Erasmus MC, and 0.90 (95% CI: 0.85–0.96) for Prague. At a score threshold of 0.15, sensitivity was 0.78, specificity 0.89, PPV 0.95, and NPV 0.59 for P+ at baseline in the pooled cohort. AI-ECG scores were higher in G+/P+ individuals at baseline than in those who developed HCM or remained P− across all sites (p<0.001; **Figure 2)**. Classification metrics at additional thresholds are presented in **Supplementary Tables S2** and **S3**.

**Figure 1.**
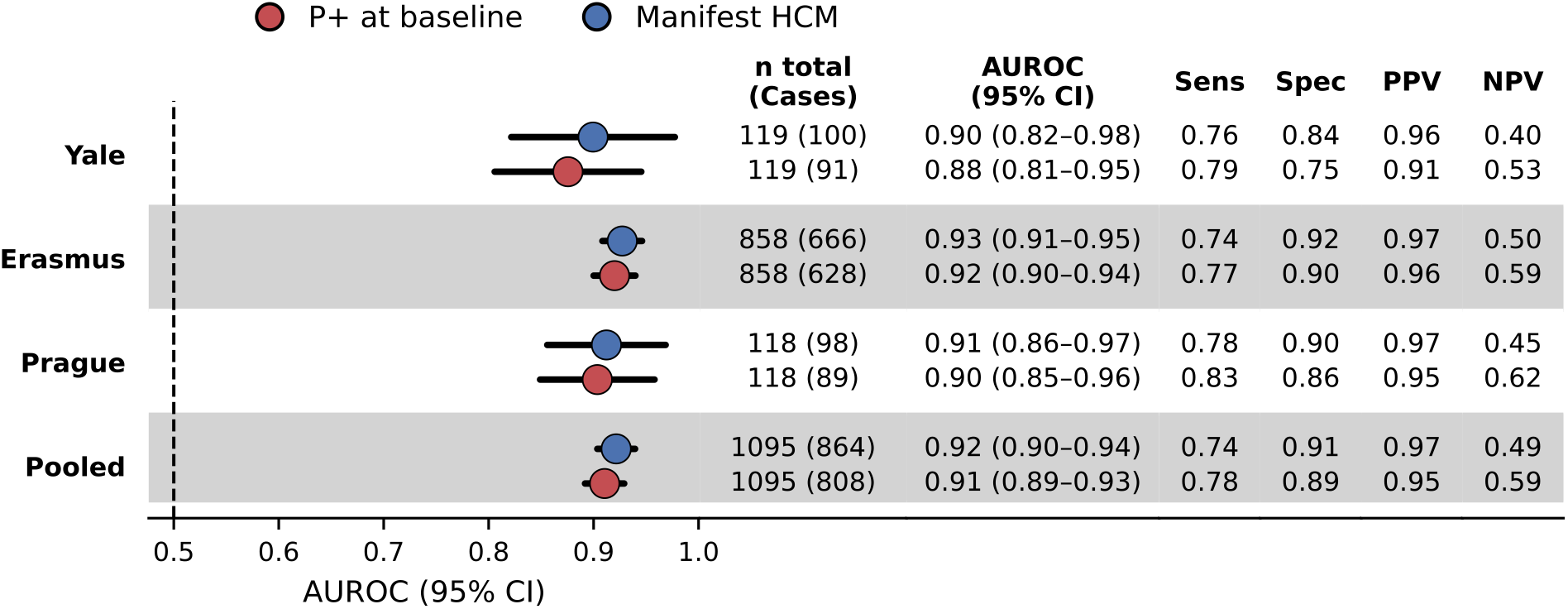
Forest plot of AUROC across cohorts. Forest plot displaying AUROC with 95% confidence intervals for P+ at baseline (red) and manifest HCM (prevalent or incident) (blue) across individual sites and the pooled cohort. Sample sizes and classification metrics at threshold 0.15 are annotated. Abbreviations: AUROC, area under the receiver operating characteristic curve; CI, confidence interval; NPV, negative predictive value; PPV, positive predictive value.

**Figure 2.**
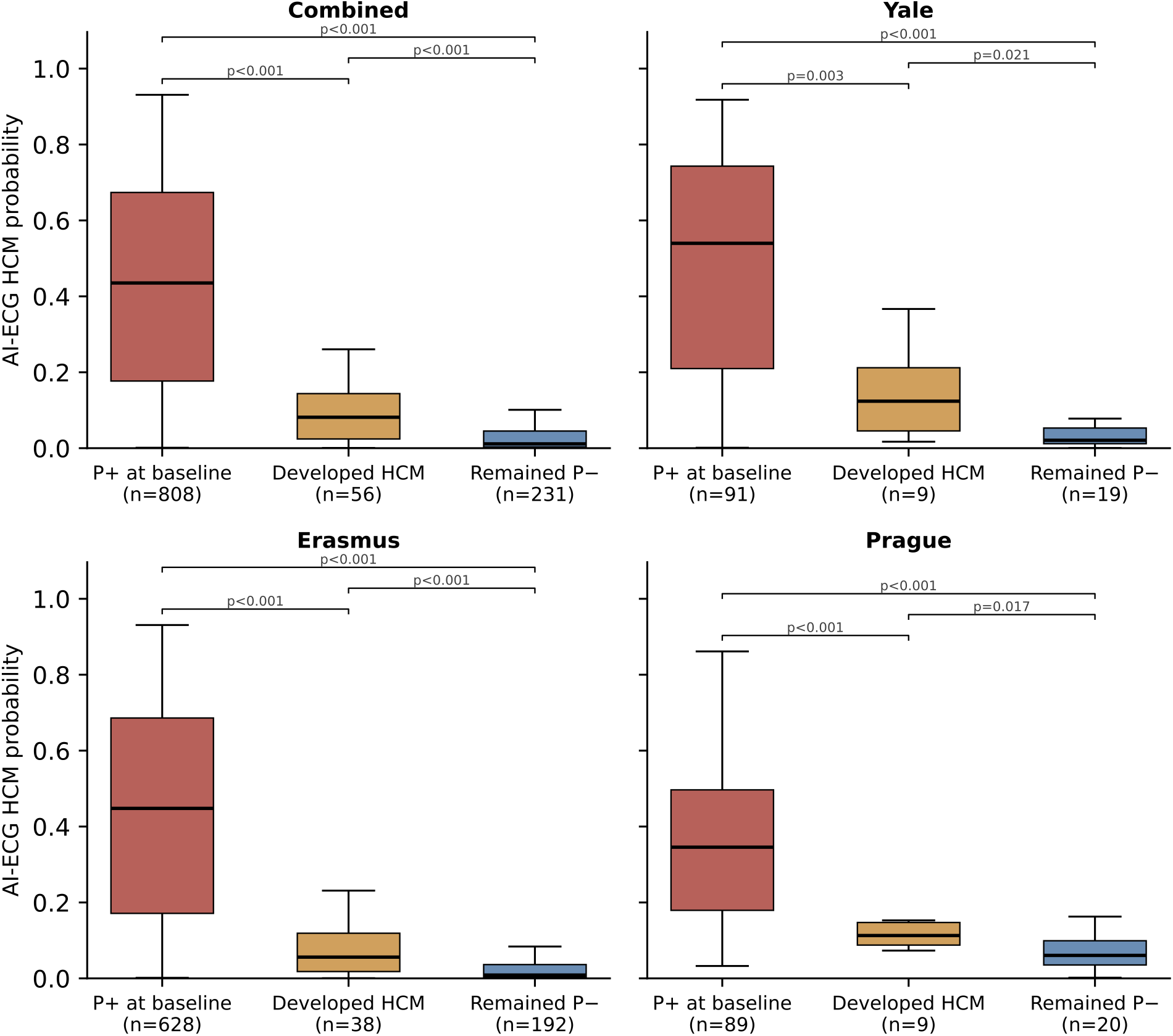
AI-ECG HCM score distribution by phenotype group. Box plots showing the distribution of AI-ECG HCM scores across three phenotype groups (P+ at baseline, Developed HCM, Remained P−) in the pooled cohort (A) and individual sites (B: Yale, C: Erasmus MC, D: Prague). Boxes indicate the median and interquartile range; whiskers extend to 1.5× IQR. P-values are derived from Mann-Whitney U tests. Sample sizes are indicated for each group. Abbreviations: AI-ECG, artificial intelligence-enhanced electrocardiography; HCM, hypertrophic cardiomyopathy.

### AI-ECG discrimination for manifest HCM

For manifest HCM, the pooled AUROC was 0.92 (95% CI: 0.90–0.94). This corresponded to a sensitivity of 0.74, specificity of 0.91, PPV of 0.97, and NPV of 0.49 at the prespecified threshold. Site-specific AUROCs were 0.90 (95% CI: 0.82–0.98) for Yale, 0.93 (95% CI: 0.91–0.95) for Erasmus MC, and 0.91 (95% CI: 0.86–0.97) for Prague **(Figure 1, Supplementary Figure S1)**.

### Prediction of HCM development during follow-up

G+/P− individuals at baseline without follow-up were excluded from both Kaplan-Meier and Cox analyses (n=48), yielding an analytic cohort of 239 individuals with a median follow-up of 6.6 years. Among these 239 individuals, 55 (23.0%) developed HCM during follow-up (**Supplementary Table S4**). Follow-up distributions by phenotype group and site are shown in **Supplementary Figure S2**. Kaplan-Meier analysis showed separation in phenotype-free survival between individuals with an index AI-ECG score ≥0.05 and those with a score <0.05 (log-rank p<0.001; 10-year phenotype-free survival 60.8% vs 85.6%; **Figure 3A**).

**Figure 3.**
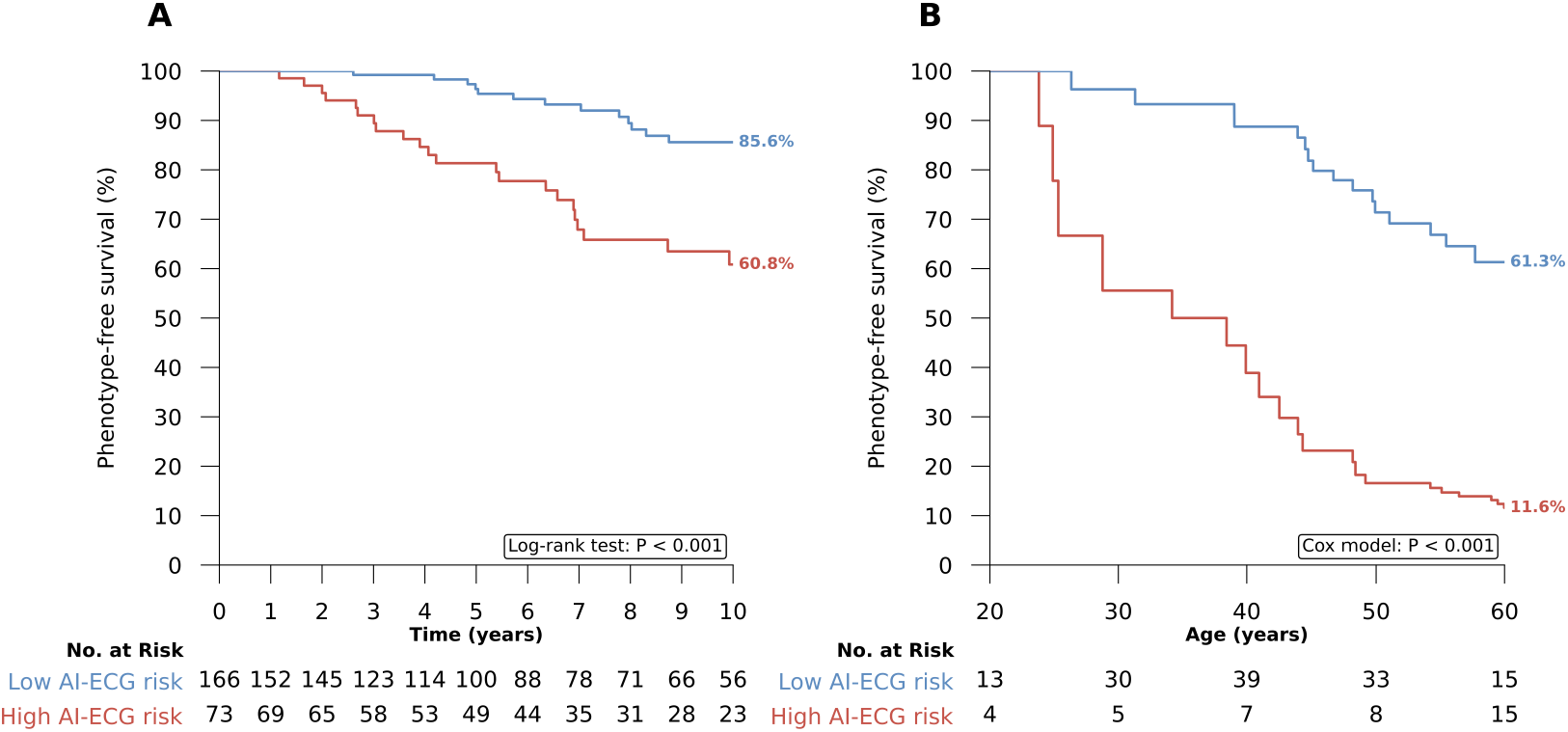
Kaplan-Meier phenotype-free survival curves by AI-ECG risk group. (A) Time from index ECG. Kaplan-Meier curves comparing phenotype-free survival between high-risk (AI-ECG score ≥0.05; red) and low-risk (AI-ECG score <0.05; blue) groups in the pooled G+/P−at baseline cohort. Follow-up is capped at 10 years. P-value is from the log-rank test. (B) Patient age as time scale. Kaplan-Meier curves with delayed entry (left truncation) at each patient’s age at index ECG, stratified by AI-ECG risk group. This analysis assesses age-dependent HCM development during follow-up. X-axis capped at age 60. P-value is from a Cox model with age as the time scale. Numbers at risk are displayed below each panel. Abbreviations: AI-ECG, artificial intelligence-enhanced electrocardiography; KM, Kaplan-Meier.

For the Kaplan–Meier curves on age as the time scale (each carrier entering follow-up at the age of their index ECG), high-risk individuals developed HCM at younger ages than low-risk individuals: estimated phenotype-free survival at age 60 was 11.6% in the high-risk group versus 61.3% in the low-risk group (Cox P < 0.001) **(Figure 3B)**. Characteristics of the 56 G+/P− individuals at baseline who developed HCM are summarized in **Supplementary Table S1**.

The AI-ECG score was associated with HCM development during follow-up, with an unadjusted HR of 1.55 (95% CI: 1.28–1.88; p<0.001) per 1-SD increase **(Table 2)**. The association persisted after adjustment for age, sex, and site (adjusted HR 1.38; 95% CI: 1.11–1.71; p=0.004), with a Harrell’s C-index of 0.73 for the pooled model **(Table 2)**. Site-specific hazard ratios were directionally consistent, with no evidence of heterogeneity across sites (I² = 0%, Cochran’s Q = 0.19, p = 0.91).

**Table 2.** Cox proportional hazards regression for HCM development during follow-up.

| Cohort | N | Events | Event<br>rate<br>(%) | Median<br>FU (y) | Unadjusted<br>HR (95% CI) | P | C-index | Adjusted HR<br>(95% CI) |
| --- | --- | --- | --- | --- | --- | --- | --- | --- |
| Yale | 18 | 9 | 50.0 | 4.5 | 1.71 (0.99–<br>2.96) | 0.054 | 0.720 | 1.38 (0.77–2.45) |
| Erasmus<br>MC | 197 | 37 | 18.8 | 7.0 | 1.51 (1.20–<br>1.90) | <0.001 | 0.679 | 1.41 (1.07–1.84) |
| Prague | 24 | 9 | 37.5 | 5.1 | 1.63 (0.76–<br>3.51) | 0.210 | 0.581 | 1.42 (0.59–3.42) |
| **Pooled** | **239** | **55** | **23.0** | **6.6** | **1.55 (1.28–**<br>**1.88)** | **<0.001** | **0.730** | **1.38 (1.11–1.71)** |

Hazard ratios (HR) are reported per 1-standard deviation increase in the standardized AI-ECG score. Adjusted models include age (standardized) and sex; the pooled model additionally adjusts for site. Abbreviations: CI, confidence interval; C-index, Harrell's concordance index; FU, follow-up.

### Subgroup analyses

Model discrimination was consistent across subgroups **(Supplementary Figures S3 and S4)**. AUROC for P+ at baseline was 0.92 (95% CI: 0.89–0.94) in males and 0.90 (0.87–0.93) in females, and 0.94 (0.92–0.96) in individuals aged <45 years versus 0.87 (0.83–0.90) in those aged ≥45 years. AUROC for manifest HCM was similarly consistent across subgroups, with point estimates ranging from 0.88 (95% CI: 0.85–0.92) in individuals aged ≥45 years to 0.94 (95% CI: 0.92–0.96) in those aged <45 years.

The prognostic association of the AI-ECG score with HCM development during follow-up was consistent across subgroups: males (HR 1.66; 95% CI: 1.30–2.12), females (HR 1.36; 95% CI: 1.02–1.83), individuals aged <45 years (HR 1.69; 95% CI: 1.35–2.10), and individuals aged ≥45 years (HR 1.49; 95% CI: 1.06–2.08). Adjustment for age and sex yielded similar results. Full subgroup results are presented in Supplementary Tables S5 and S6. A sensitivity analysis using age 40 as an alternative cutoff yielded similar results **(Supplementary Table S7)**.

AI-ECG discrimination was consistent across gene groups **(Supplementary Table S8)**. For P+ at baseline, AUROC was 0.92 (95% CI: 0.90–0.94) for MYBPC3 (n=751), 0.89 (95% CI: 0.82–0.96) for MYH7 (n=163), and 0.89 (95% CI: 0.84–0.94) for other gene variants (n=181). For manifest HCM (prevalent or incident) phenotype, the corresponding AUROCs were 0.91 (95% CI: 0.89–0.94), 0.95 (0.91–1.00), and 0.93 (0.89–0.97), respectively.

### Study population for polygenic risk analyses

We identified 57,007 unrelated White British UK Biobank participants (mean age 55.0 ± 7.5 years; 50.9% female) with available 12-lead ECG recordings and whole-genome sequencing data. Prevalent comorbidities included hypertension (22.0%), hypercholesterolemia (11.2%), coronary artery disease (7.5%), and type 2 diabetes (4.1%), and participants had a mean BMI of 26.6 ± 4.2 kg/m². Among 57,007 participants, 51 had a diagnosis of HCM **(Table 3)**.

**Table 3.**
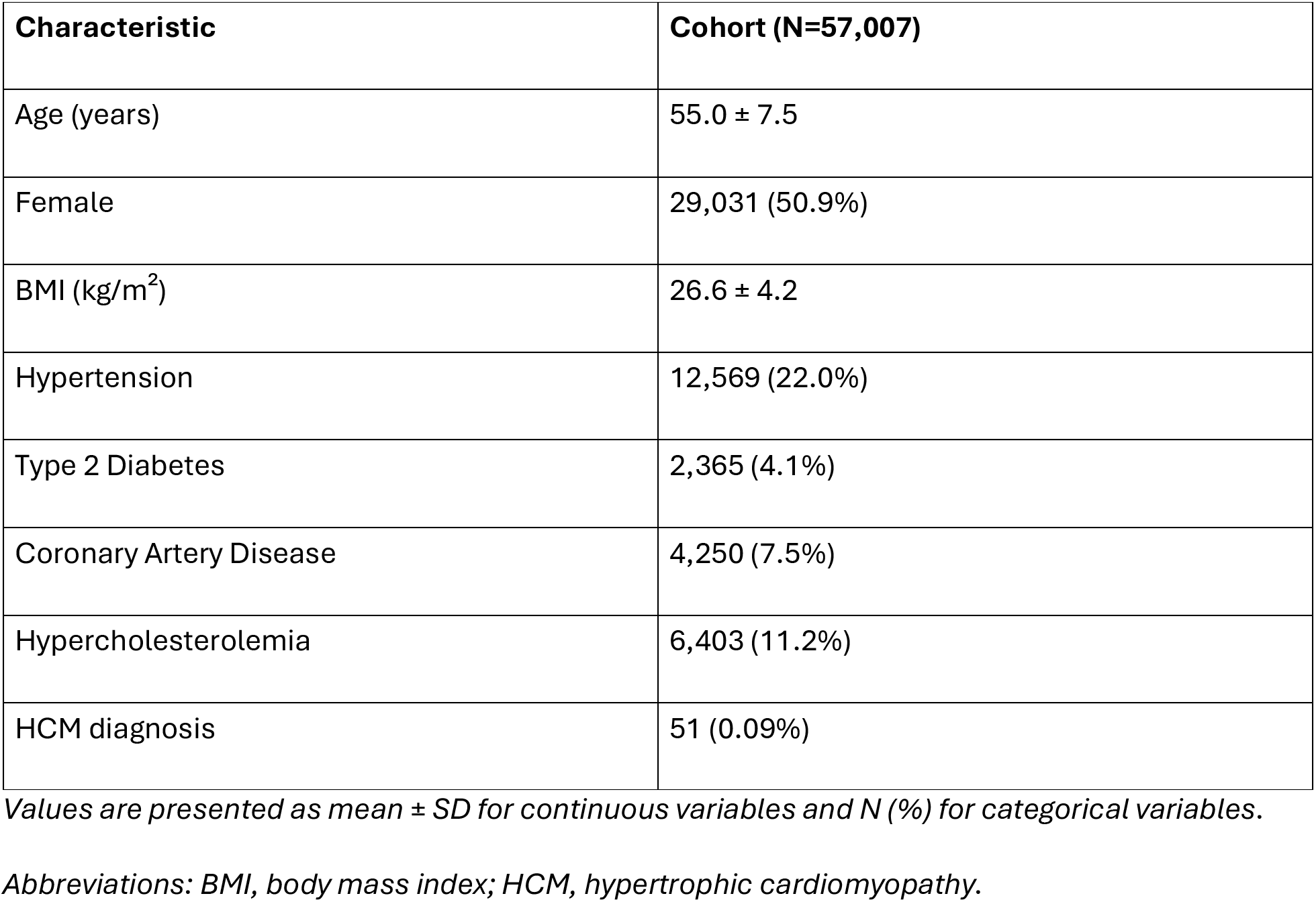
Baseline characteristics of UK Biobank participants (N=57,007).

### Complementary value of AI-ECG and polygenic risk in the UK Biobank

Among the included UK Biobank participants, we calculated PRS and AI-ECG scores (Table 3). HCM prevalence increased across the four risk strata: 0.02% (9/41,721) in the both-low group, 0.09% (9/10,255) in PRS-only high, 0.41% (16/3,883) in AI-ECG-only high, and 1.48% (17/1,148) in the group where both AI-ECG and PRS were high **(Figure 4A)**. After adjustment for age and sex, the odds of HCM were 60.2-fold higher in individuals with both a high PRS and a high AI-ECG score than in the reference group with low PRS and low AI-ECG score (95% CI: 26.5–137.2). By comparison, the odds ratios were 15.0 (95% CI: 6.5–34.7) for high AI-ECG score alone and 4.1 (95% CI: 1.6–10.4) for high PRS alone **(Figure 4B, Supplementary Table S9)**. There was no significant interaction between the AI-ECG score and PRS (p=0.69), consistent with additive risk on the log-odds scale.

**Figure 4.**
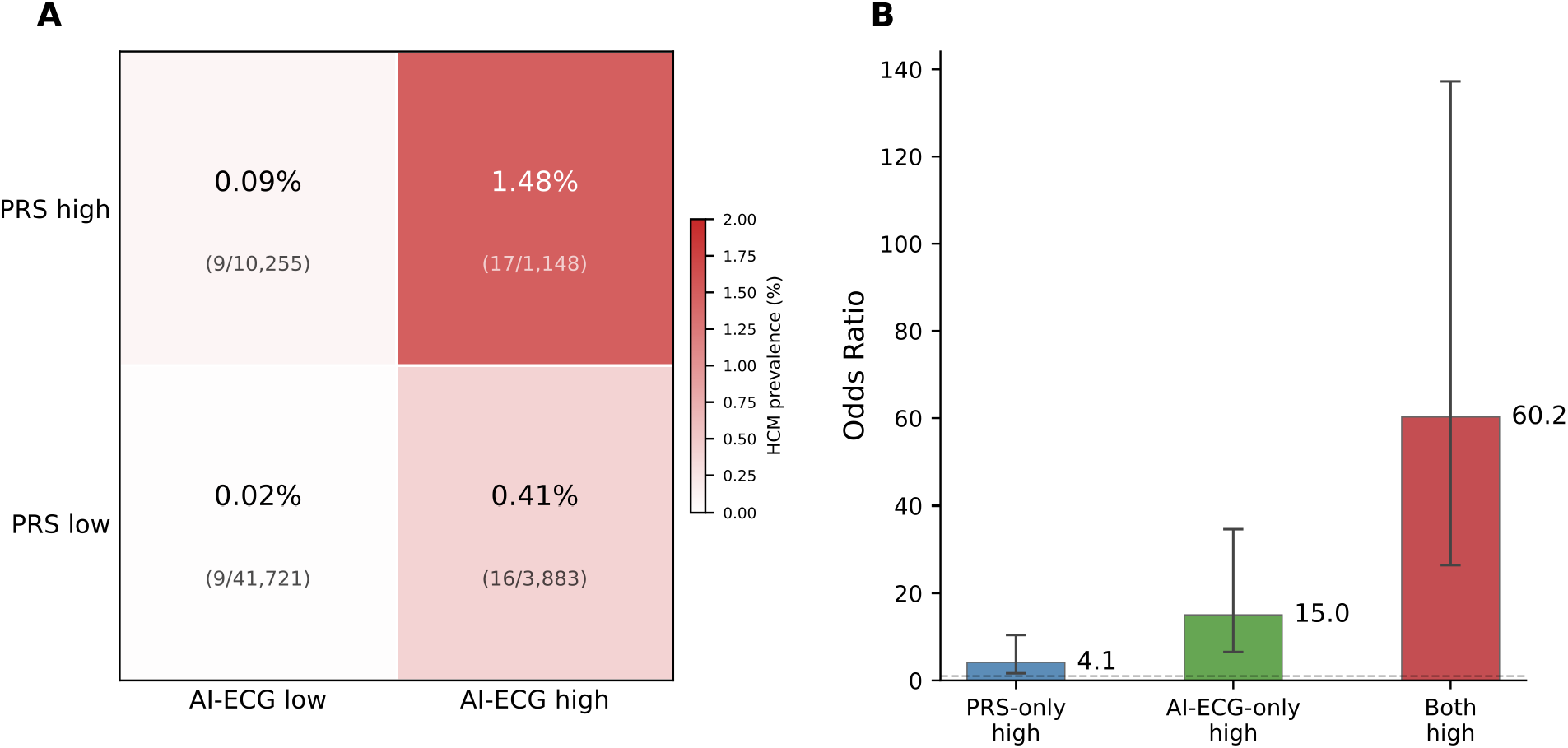
Complementarity of AI-ECG and polygenic risk score for HCM in the UK Biobank. (A) Heatmap showing HCM prevalence across four risk strata defined by AI-ECG score (high: ≥0.15; low: <0.15) and polygenic risk score (PRS; high: ≥80th percentile; low: <80th percentile) in 57,007 UK Biobank participants. Cell values show prevalence and case counts. Color intensity reflects prevalence. (B) Odds ratios for HCM in each risk stratum compared with the both-low reference group. HCM cases were observed across all four strata, consistent with information from complementary channels. Abbreviations: AI-ECG, artificial intelligence-enhanced electrocardiography; HCM, hypertrophic cardiomyopathy; OR, odds ratio; PRS, polygenic risk score.

## Discussion

In this study spanning three international centers and a large prospective cohort, we found that a validated AI-ECG model identified the baseline HCM phenotype in carriers of pathogenic sarcomere variants, and refined risk stratification among individuals with elevated polygenic risk. AI-ECG applied to widely available ECG images or signals identified the HCM phenotype at baseline (P+) with strong discrimination. Among G+/P− individuals at baseline, AI-ECG identified those at higher risk of developing HCM. In a secondary analysis of the UK Biobank, the AI-ECG HCM score captured information complementary to polygenic risk; individuals with both elevated genetic risk and a high AI-ECG score had 60-fold higher odds of HCM than those with low polygenic risk and a low AI-ECG score. Together, these findings support AI-ECG as a readily deployable tool for risk stratification in a high-risk population in which current surveillance relies on serial imaging without individualized risk assessment.

These findings add to the evidence supporting refined risk stratification in individuals at risk of HCM. Cascade screening of relatives is the standard of care and has identified a growing population of sarcomere variant carriers without overt HCM (G+/P−) who enter lifelong imaging surveillance.^5,11^ In a substantial proportion of clinically diagnosed HCM cases, no causal sarcomere variant is identified; yet first-degree relatives remain at elevated risk and are recommended to undergo clinical screening, further expanding the at-risk population.^3,22,23^ The data underpinning indefinite surveillance in these groups remain limited, and the penetrance of pathogenic sarcomere variants is low; many individuals undergo years of imaging without HCM development during follow-up.^24–26^ Pre-clinical HCM involves subtle myocardial structural changes, and electrocardiographic abnormalities often precede overt left ventricular hypertrophy.^2,6,27^ Yet most carriers show no overt changes at initial evaluation, leaving clinicians with few accessible tools to prioritize surveillance among a growing at-risk population.^2,28^ Prior reports have demonstrated that AI-ECG extracts features from a ubiquitous, low-cost test that enable phenotype detection beyond expert reader interpretation, providing a scalable approach to HCM detection.^16,18^ However, deployment in unselected populations is constrained by low PPV given low disease prevalence.^18^ Our findings extend this work by evaluating AI-ECG as a first-line risk-stratification tool in a genetically at-risk population with substantially higher baseline prevalence. This approach concentrates screening where it is most likely to yield actionable findings.

Our findings further indicate that AI-ECG may serve as an accessible phenotypic marker that complements polygenic risk and may have implications for clinical practice. Beyond rare pathogenic variants, PRS captures the polygenic architecture of HCM, modifying penetrance among carriers and stratifying risk in the general population.^7,19,29^ Although PRS improve risk stratification for HCM, they reflect inherited susceptibility rather than current phenotypic state. They have been derived almost exclusively in European-ancestry populations with limited generalizability, require specialized genotyping infrastructure not available at the point of care, and offer modest discrimination on their own.^30^ In contrast, AI-ECG analysis of standard ECG images is easy to implement, generalizes across demographic subgroups, and provides a repeatable marker of early disease expression. Used alongside imaging-based screening, it may support a more individualized strategy in which monitoring intensity is guided by near-term risk.^16,31^

As a standalone tool in carriers of pathogenic sarcomere variants, AI-ECG may serve two complementary functions: detecting overt and subclinical phenotypes in relatives undergoing cascade screening, and identifying G+/P− individuals at baseline who are at higher risk of development of HCM. Beyond standalone use, AI-ECG can be combined with PRS to simultaneously capture germline susceptibility and early phenotypic expression. This combined approach focuses screening on those most likely to harbor early disease, improving the yield of risk stratification. However, whether a negative AI-ECG result can safely defer imaging requires prospective evaluation and should not be inferred from the current study. Together, these findings support AI-ECG as both a standalone risk-stratification tool in genetically at-risk cohorts and as a complementary marker that, alongside polygenic risk, enables individualized phenotype prediction.

Strengths of this study include multicenter validation in a large sample across three independent international sites, use of a previously validated AI-ECG model with a pre-specified threshold, the ability to leverage widely available ECG images, and consistent performance across sites, sexes, age groups, and gene variants. Several limitations should be acknowledged. First, the study is retrospective in design, and prospective validation is needed to confirm the clinical utility of AI-ECG-guided surveillance. Second, the relatively small number of cases who developed HCM during follow-up limited statistical power for site-specific and gene-stratified subgroup analyses. Third, index ECG selection differed across sites. Yale used a 90-day window around imaging, whereas Erasmus MC and Motol University Hospital used the first available ECG, which may explain differences in site-specific discrimination. Fourth, we did not distinguish between pathogenic and likely pathogenic variants, which may differ in their associated risk of HCM development. Fifth, the sarcomere variant cohorts were drawn from two European and one North American center, and performance of AI-ECG in more diverse ancestral populations remains to be established. Sixth, the UK Biobank is a volunteer-based cohort that is healthier and predominantly of European ancestry, and the small absolute number of HCM cases limited the precision of stratum-specific risk estimates. Despite these limitations, this is, to our knowledge, among the largest multicenter evaluations of risk stratification in sarcomere variant carriers and the first to demonstrate complementarity between an image-based AI-ECG phenotypic marker and polygenic risk for HCM, supporting prospective evaluation of genetically informed risk stratification.

## Conclusions

A validated image-based AI-ECG model identified the HCM phenotype at baseline and predicted development of HCM during follow-up in carriers of pathogenic sarcomere variants across three international centers. In a population-based analysis, AI-ECG and polygenic risk score were independent and additive markers of HCM risk, with their combination identifying individuals at markedly elevated odds of disease. Together, these findings support AI-ECG as a scalable, low-cost tool for individualized HCM risk stratification across both monogenic and polygenic at-risk populations.

## Data Availability

The data is not available due to privacy concerns

## Non-standard abbreviations and acronyms

AI: Artificial Intelligence
AI-ECG: Artificial Intelligence-Enhanced Electrocardiogram
AUROC: Area Under the Receiver Operating Characteristic Curve
CMR: Cardiac Magnetic Resonance Imaging
CNN: Convolutional Neural Network
ECG: Electrocardiogram
G+: Genotype-Positive
HCM: Hypertrophic Cardiomyopathy
LLM: Large Language Model
P+: Phenotype-Positive
P−: Phenotype-Negative
PRS: Polygenic Risk Score
UKB: UK Bioban

