## Supplements for "Artificial Intelligence-Enhanced Electrocardiography for Detection and Prediction of Hypertrophic Cardiomyopathy across Monogenic and Polygenic Susceptibility"

**Supplementary Materials**

**Artificial Intelligence-Enhanced Electrocardiography for Hypertrophic Cardiomyopathy Risk Stratification in Individuals at Monogenic or Polygenic Risk**

Philip M. Croon, MD, Rohan Khera, MD, MS

### SUPPLEMENTARY METHODS

**LLM Phenotype Annotation Prompt**

The following prompt template was used with Qwen 32B (deployed locally via Ollama) for automated phenotype annotation of echocardiography and CMR reports at the Yale site:

You are a clinical cardiology AI assistant analyzing cardiac imaging reports for a research study on hypertrophic cardiomyopathy (HCM). Your task is to determine whether a given imaging report describes findings consistent with HCM phenotype expression.

DEFINITION OF HCM PHENOTYPE:

- Left ventricular hypertrophy (LVH) with wall thickness >= 13mm in any segment, OR

- Asymmetric septal hypertrophy (ASH), OR

- Systolic anterior motion (SAM) of the mitral valve, OR

- Left ventricular outflow tract (LVOT) obstruction, OR

- Explicit mention of HCM or hypertrophic cardiomyopathy diagnosis

IMPORTANT NOTES:

- Concentric LVH alone may not indicate HCM. Look for asymmetry or other HCM features.

- Wall thickness of 13-14mm is borderline; if the report attributes it to hypertension alone, classify as NOT HCM.

- If the report mentions "cannot rule out HCM" or "possible HCM", classify as NOT HCM (uncertain).

- Reports that mention prior HCM diagnosis or history of HCM should be classified as HCM POSITIVE.

OUTPUT FORMAT:

Respond with ONLY a JSON object:

{"phenotype": "positive" or "negative", "confidence": "high" or "medium" or "low", "reasoning": "brief explanation"}

IMAGING REPORT:

{report_text}

**SUPPLEMENTARY TABLES**

Supplementary Table S1. Baseline characteristics by phenotype group. Characteristics are presented for the three phenotype groups in the pooled cohort (P+ at baseline, Developed HCM, Remained P−) and the overall cohort. Continuous variables are reported as median (interquartile range); categorical variables as count (percentage).

| Characteristic | P+ at baseline (n=808) | Developed HCM (n=56) | Remained P− (n=231) | Total (n=1,095) |
| --- | --- | --- | --- | --- |
| Age, years, median (IQR) | 47.7 (36.0–58.0) | 45.8 (31.8–55.0) | 40.0 (28.1–49.5) | 46.0 (34.0–56.0) |
| Female, n (%) | 455 (56.3%) | 24 (42.9%) | 91 (39.4%) | 570 (52.1%) |
| Male, n (%) | 353 (43.7%) | 32 (57.1%) | 140 (60.6%) | 525 (47.9%) |
| Gene, n (%) |  |  |  |  |
| MYBPC3 | 541 (67.0%) | 41 (73.2%) | 169 (73.2%) | 751 (68.6%) |
| MYH7 | 128 (15.8%) | 7 (12.5%) | 28 (12.1%) | 163 (14.9%) |
| MYL2 | 33 (4.1%) | 1 (1.8%) | 3 (1.3%) | 37 (3.4%) |
| MYL3 | 8 (1.0%) | — | 1 (0.4%) | 9 (0.8%) |
| TNNI3 | 21 (2.6%) | 2 (3.6%) | 4 (1.7%) | 27 (2.5%) |
| TNNT2 | 31 (3.8%) | — | 9 (3.9%) | 40 (3.7%) |
| TPM1 | 5 (0.6%) | 1 (1.8%) | — | 6 (0.5%) |
| ACTN2 | 3 (0.4%) | 1 (1.8%) | 2 (0.9%) | 6 (0.5%) |
| Unknown/Other | 38 (4.7%) | 3 (5.4%) | 15 (6.5%) | 56 (5.1%) |
| AI-ECG score, median (IQR) | 0.4 (0.2–0.7) | 0.1 (0.0–0.1) | 0.0 (0.0–0.0) | 0.3 (0.0–0.6) |

Supplementary Table S2. Classification metrics at multiple AI-ECG score thresholds for detection of P+ at baseline in the pooled cohort.

| **Threshold** | **Sensitivity** | **Specificity** | **PPV** | **NPV** | **N Predicted +** | **N True +** |
| --- | --- | --- | --- | --- | --- | --- |
| 0.05 | 0.91 | 0.70 | 0.90 | 0.74 | 821 | 736 |
| 0.10 | 0.84 | 0.83 | 0.93 | 0.64 | 726 | 676 |
| 0.15 | 0.78 | 0.89 | 0.95 | 0.59 | 663 | 630 |
| 0.20 | 0.73 | 0.92 | 0.96 | 0.55 | 611 | 589 |
| 0.25 | 0.67 | 0.95 | 0.97 | 0.51 | 557 | 542 |
| 0.30 | 0.63 | 0.95 | 0.97 | 0.48 | 524 | 510 |
| 0.50 | 0.43 | 0.98 | 0.98 | 0.38 | 352 | 346 |

Supplementary Table S3. Classification metrics at multiple AI-ECG score thresholds for detection of manifest HCM (prevalent or incident) in the pooled cohort.

| **Threshold** | **Sensitivity** | **Specificity** | **PPV** | **NPV** | **N Predicted +** | **N True +** |
| --- | --- | --- | --- | --- | --- | --- |
| 0.05 | 0.89 | 0.78 | 0.94 | 0.66 | 821 | 771 |
| 0.10 | 0.81 | 0.88 | 0.96 | 0.55 | 726 | 699 |
| 0.15 | 0.74 | 0.91 | 0.97 | 0.49 | 663 | 643 |
| 0.20 | 0.69 | 0.95 | 0.98 | 0.45 | 611 | 599 |
| 0.25 | 0.64 | 0.97 | 0.99 | 0.42 | 557 | 550 |
| 0.30 | 0.60 | 0.97 | 0.99 | 0.39 | 524 | 517 |
| 0.50 | 0.40 | 0.99 | 0.99 | 0.31 | 352 | 349 |

Supplementary Table S4. Survival cohort characteristics.

| Site | N | Events | Event rate (%) | Median follow-up (years) |
| --- | --- | --- | --- | --- |
| Yale | 18 | 9 | 50.0 | 4.5 |
| Erasmus MC | 197 | 37 | 18.8 | 7.0 |
| Prague | 24 | 9 | 37.5 | 5.1 |
| Pooled | 239 | 55 | 23.0 | 6.6 |

*The survival cohort includes all G+/P− individuals at baseline (Developed HCM + Remained P−). Patients with zero or negative follow-up time (n=48; 47 with zero and 1 with negative follow-up time) were excluded from Cox regression analyses, resulting in an analytic sample of N=239 with 55 events for those models. FU = follow-up; IQR = interquartile range.*

Supplementary Table S5. Subgroup discrimination (AUROC) by sex and age group in the pooled cohort.

| **Subgroup** | **N** | **AUROC P+ at baseline (95% CI)** | **AUROC manifest HCM (95% CI)** |
| --- | --- | --- | --- |
| Male | 525 | 0.92 (0.89–0.94) | 0.93 (0.91–0.95) |
| Female | 570 | 0.90 (0.87–0.93) | 0.91 (0.88–0.94) |
| <45 years | 506 | 0.94 (0.92–0.96) | 0.94 (0.92–0.96) |
| ≥45 years | 589 | 0.87 (0.83–0.90) | 0.88 (0.85–0.92) |
| Overall | 1,095 | 0.91 (0.89–0.93) | 0.92 (0.90–0.94) |

Supplementary Table S6. Subgroup hazard ratios for development of HCM during follow-up by sex and age group in the pooled cohort. The Overall row is adjusted for age and sex only; the site-adjusted pooled estimate is reported in Table 2 (unadjusted HR 1.55, 95% CI 1.28–1.88; adjusted HR 1.38, 95% CI 1.11–1.71). N denotes the number of individuals in each subgroup of the full cohort; events occurred among G+/P− individuals at baseline with available follow-up.

| **Subgroup** | **N** | **Events** | **Unadjusted HR (95% CI)** | **P** | **Adjusted HR (95% CI)** | **P** |
| --- | --- | --- | --- | --- | --- | --- |
| Male | 525 | 32 | 1.66 (1.30–2.12) | <0.001 | 1.45 (1.11–1.89) | 0.006 |
| Female | 570 | 23 | 1.36 (1.02–1.83) | 0.037 | 1.30 (0.91–1.84) | 0.150 |
| <45 years | 506 | 26 | 1.69 (1.35–2.10) | <0.001 | 1.69 (1.34–2.12) | <0.001 |
| ≥45 years | 589 | 29 | 1.49 (1.06–2.08) | 0.022 | 1.36 (0.95–1.94) | 0.088 |
| Overall | 1,095 | 55 | 1.56 (1.30–1.88) | <0.001 | 1.42 (1.16–1.75) | <0.001 |

Supplementary Table S7. Age cutoff sensitivity analysis. Comparison of age-stratified subgroup analyses using age 40 versus age 45 as the stratification cutoff. AUROC values are presented for detection of P+ at baseline and manifest HCM. Classification metrics are reported at the standard threshold of 0.15.

| **Outcome** | **Age subgroup** | **N** | **AUROC** | **AUROC 95% CI** | **Sensitivity** | **Specificity** | **PPV** | **NPV** |
| --- | --- | --- | --- | --- | --- | --- | --- | --- |
| P+ at baseline | <40 | 391 | 0.940 | 0.918–0.962 | 0.922 | 0.787 | 0.890 | 0.843 |
| P+ at baseline | ≥40 | 704 | 0.882 | 0.852–0.912 | 0.906 | 0.629 | 0.899 | 0.646 |
| P+ at baseline | <45 | 506 | 0.938 | 0.917–0.958 | 0.911 | 0.800 | 0.900 | 0.819 |
| P+ at baseline | ≥45 | 589 | 0.868 | 0.833–0.902 | 0.911 | 0.564 | 0.894 | 0.611 |
| Manifest HCM | <40 | 391 | 0.938 | 0.915–0.960 | 0.892 | 0.851 | 0.936 | 0.764 |
| Manifest HCM | ≥40 | 704 | 0.901 | 0.872–0.930 | 0.893 | 0.718 | 0.941 | 0.571 |
| Manifest HCM | <45 | 506 | 0.940 | 0.920–0.959 | 0.882 | 0.860 | 0.941 | 0.741 |
| Manifest HCM | ≥45 | 589 | 0.884 | 0.848–0.920 | 0.900 | 0.659 | 0.938 | 0.537 |

Supplementary Table S8. Gene-stratified AUROC.

| **Gene** | **N** | **AUROC P+ at baseline (95% CI)** | **AUROC manifest HCM (95% CI)** |
| --- | --- | --- | --- |
| MYBPC3 | 751 | 0.92 (0.90–0.94) | 0.91 (0.89–0.94) |
| MYH7 | 163 | 0.89 (0.82–0.96) | 0.95 (0.91–1.00) |
| Other | 181 | 0.89 (0.84–0.94) | 0.93 (0.89–0.97) |

*AUROC = area under the receiver operating characteristic curve; CI = confidence interval.*

Supplementary Table S9. HCM prevalence and odds ratios by combined AI-ECG and PRS risk strata in the UK Biobank.

| Group | N | HCM cases | Prevalence (%) | Unadjusted OR (95% CI) | Adjusted OR (95% CI)* |
| --- | --- | --- | --- | --- | --- |
| Both low | 41,721 | 9 | 0.02 | Ref | Ref |
| PRS-only high | 10,255 | 9 | 0.09 | 4.07 (1.62–10.26) | 4.14 (1.64–10.43) |
| AI-ECG-only high | 3,883 | 16 | 0.41 | 19.18 (8.47–43.42) | 15.05 (6.52–34.71) |
| Both high | 1,148 | 17 | 1.48 | 69.66 (30.99–156.61) | 60.25 (26.45–137.23) |

Risk strata defined by AI-ECG score (high: ≥0.15; low: <0.15) and PRS (high: ≥80th percentile; low: <80th percentile). Reference group: both low. Adjusted odds ratios were estimated from logistic regression adjusted for age and sex.

### Supplementary Figure Legends

### Supplementary Figure S1. Receiver operating characteristic curves for detection of P+ at baseline and manifest HCM.

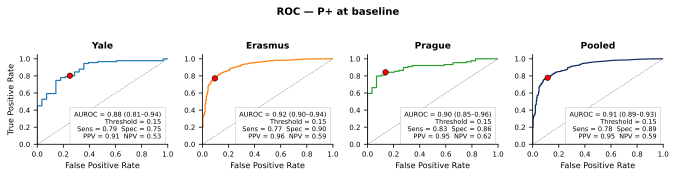

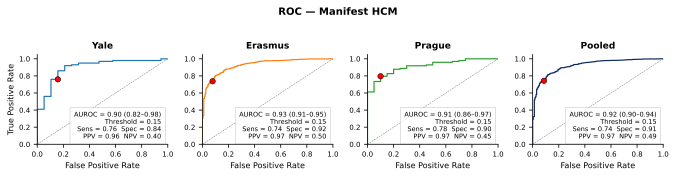

ROC curves for the AI-ECG model's discrimination of P+ at baseline (top row) and manifest HCM (bottom row) across individual sites (Yale, Erasmus MC, Prague) and the pooled cohort. AUROC values with 95% confidence intervals (DeLong's method) are shown for each curve. Red markers indicate the prespecified threshold of 0.15, with corresponding sensitivity, specificity, positive predictive value, and negative predictive value annotated. Abbreviations: AI-ECG, artificial intelligence-enhanced electrocardiography; AUROC, area under the receiver operating characteristic curve; HCM, hypertrophic cardiomyopathy; NPV, negative predictive value; PPV, positive predictive value; ROC, receiver operating characteristic.

Supplementary Figure S2. Follow-up distribution by phenotype group and site.

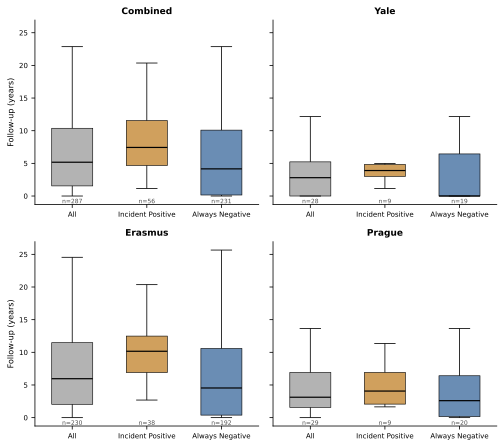

Box plots showing follow-up duration (years) by phenotype group (All, Developed HCM, Remained P−) across sites and pooled. Boxes indicate median and interquartile range; whiskers extend to 1.5× IQR. Sample sizes are indicated below each group. Abbreviations: IQR, interquartile range.

Supplementary Figure S3. Forest plot of AUROC by sex and age group.

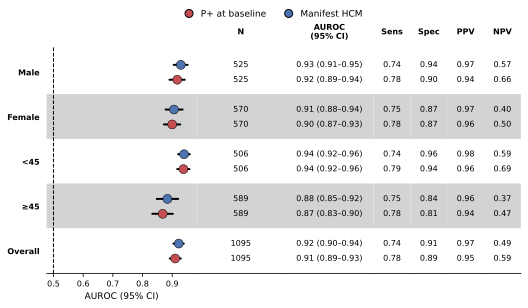

Forest plot of AUROC stratified by sex and age group for detection of P+ at baseline. Abbreviations: AI-ECG, artificial intelligence-enhanced electrocardiography; AUROC, area under the receiver operating characteristic curve; CI, confidence interval.

Supplementary Figure S4. Forest plot of hazard ratios by sex and age group.

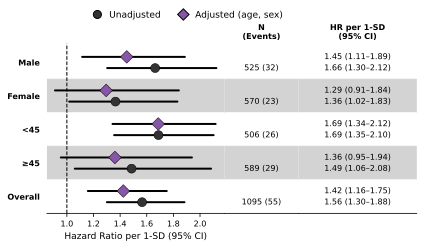

Forest plot of hazard ratios per 1-SD increase in AI-ECG score stratified by sex and age group, showing unadjusted (circles) and adjusted (diamonds; adjusted for age and sex) estimates with 95% confidence intervals. Abbreviations: AI-ECG, artificial intelligence-enhanced electrocardiography; CI, confidence interval; HR, hazard ratio; SD, standard deviatio
